# Detecting Incipient Heart Failure in Asymptomatic Patients with Normal Ejection Fraction and comparisons with patients with heart failure and preserved ejection fraction using TimeSformer for classifying Echocardiography videos

**DOI:** 10.1101/2024.10.22.24315954

**Authors:** Srilakshmi M Adhyapak, Prahlad G Menon

**Author notes:** Address for Correspondence: Srilakshmi M Adhyapak, Prof and Head, Dept of Cardiology, St. John’s Medical College Hospital, Bangalore 560034, India.

## Abstract

**Background:** Recently deep learning models have helped differentiate echocardiography images of patients with heart failure with preserved ejection fraction (HFpEF) from normal controls. Our aim was to develop a model capable of detecting early signs of heart failure in asymptomatic patients with a normal ejection fraction.

**Methods:** We employed the TimeSformer, a video transformer model that classifies video data using a novel attentionbased mechanism. This self-attention mechanism that diverges from traditional convolutional neural networks (CNNs). It focuses on relevant parts of the video across both space and time, split into spatial attention, which processes each frame individually, and temporal attention, which integrates information across different frames. The training and validation of the TimeSformer model were conducted on the same dataset of echocardiography videos from 50 normal controls and 80 patients diagnosed with HFpEF, employing 5-fold cross-validation to ensure robust performance evaluation.

**Results:** The TimeSformer model effectively identified HFpEF in patients, as all diagnosed with HFpEF were flagged as abnormal. The echocardiography assessment, along with NT pro BNP levels, supported the diagnoses, with patients showing NT pro BNP levels of 1016±32 pg/ml. Conversely, 26 out of 50 normal controls were correctly identified. While 24 normal controls were identified as abnormal. Their NT pro BNP levels were 52±12 pg/ml. At 2 years 8 (33.3%) controls identified as abnormal developed symptoms of heart failure with NT pro BNP levels of 800±41 pg/ml.

**Conclusions:** The TimeSformer model demonstrated capability in identifying subtle deviations indicative of incipient heart failure in videos of normal controls, despite normal NT pro BNP levels. A significant number of these controls developed heart failure with elevated NT pro BNP levels at 2 years.

## Introduction

Heart failure with preserved ejection fraction (HFpEF) constitutes a clinical spectrum which constitutes a diagnostic dilemma as well as a therapeutic challenge in treating it[1-5].

HFpEF is a heterogenous syndrome associated with various comorbidities, wherein cardiac and noncardiac factors contribute to elevated intracardiac filling pressure, resulting in signs and symptoms of HF[4]. There is large intra-observer and inter-observer variabilities in the echocardiographic evaluation of this entity. Especially Doppler assessment of LV filling pressures has great inter-observer and intra-observer variability. Recent work in artificial intelligence (AI) computer vision techniques offer great promise of better interpretation of various information in medical data, especially images. Recent AI studies have combined clinical parameters and manual echocardiographic measurements to classify diastolic dysfunction and HFpEF. Development of an approach using an echocardiographic video might obviate the need for complex Doppler assessment.

Recently, an AI model was used to automatically detect HFpEF by only using the apical 4-chamber (A4C) TTE video clip. This view was selected because it includes much information (chamber sizes, wall thicknesses, annulus motion, etc) and is routinely acquired in imaging protocols. This novel AI HFpEF model provided fewer nondiagnostic outputs than current clinical scores and identified patients with worse survival [6].

We hypothesized that an AI model based on the A4C TTE videoclip could identify subtle abnormalities in normal individuals who might have subclinical disease undistinguishable by routine clinical scores and biomarkers.

## Methods

This was a prospective study done in the department of cardiology at St. John’s Medical College Hospital, Bangalore; India. The study was approved by the Institutional Ethics Committee Study No: 289/2023. Written consent for use of TTE analysis and relevant clinical patient information was exempted by the participating Institutional Review Boards due to the use of deidentified data.

## Model training and validation

All patients presenting to the Outpatient/ In-patient with symptoms of heart failure were studied. They all underwent a comprehensive TTE examination by trained echocardiographers whose inter-observer and intra-observer variability was not significant. Patients were included based on preserved ejection fraction and increased intracardiac filling pressures, which constituted patients with HFpEF. Controls were randomly selected in distribution with the patients with respect to age, sex and year of echocardiographic examination.

### Clinical Diagnosis of heart failure

Clinical diagnosis of HF, based on the ACC/AHA guidelines for diagnosis of heart failure, (case) or lack of this diagnosis (control) was studied. All patients had NT pro-BNP levels in addition to the routine blood panels. The controls also had NT pro BNP levels documented.

### Overview of the AI model

#### AI model outputs

##### Comparison of AI model with clinical practice

To test the hypothesis that the classification accuracy of the developed AI HFpEF model, based on analysis of a single A4C video clip, was acceptable, we compared observed sensitivity and specificity in the data set to average reported data in the literature (sensitivity, 74%; specificity, 65%) [11].

The TimeSformer is a convolution-free approach to video classification built exclusively on self-attention over space and time. It adapts the standard Transformer architecture to video by enabling spatiotemporal feature learning directly from a sequence of frame level patches.

Here temporal attention and spatial attention are separately applied within each block, leading to a very good video classification accuracy. When compared to 3D convolutional networks, this model is faster to train, it can achieve dramatically higher test efficiency, and can be applied to much longer video clips (over one minute long)[15].

##### Statistical methods

The sensitivity and specificity of AI model was estimated against the clinical diagnosis. The positive and negative predictive values, negative and positive likelihood ratios with 95% Confidence interval is also reported. Categorical variables were summarized as numbers (%) and continuous variables as mean (SD). The normal distribution assumption of continuous variables was examined using Q-Q plot. The association of AI with categorical variables was examined using Chi-square test and with continuous variables using independent sample t test. The Kaplan Meir plot of time to heart failure in the 8 control subjects who developed heart failure at later time is also presented. Statistical significance was considered at 5% level. All analyses were carried out using STATA release 15 (StataCorp, College Station, Tex).

## Results

### Classification accuracy of the AI model

The final model training and validation data set comprised 130 patients (80 cases, 50 controls) with 130 A4C video clips (80 cases, and 50 controls). Classification performance in the training and validation data sets was high corresponding to a sensitivity and specificity of 100% and 61.9%, respectively. These values corresponded to a positive predictive value of 84.62% (95% CI: 78.91% to 89.00%) and a negative predictive value of 100.00% (95% CI: 86.77% to 100.00%) respectively. The accuracy being 87.69% (95% CI: 80.78% to 92.80%).

### Reproducibility of the AI model

The model demonstrated good agreement for repeatability of all model outputs. The model demonstrated perfect agreement for negative likelihood ratio of diagnostic outputs (0.00), positive likelihood ratio of diagnostic outputs (2.62; 95% CI: 1.79 to 3.86).

### Accuracy of AI model in comparison with clinical scores for HFpEF

To assess whether the AI model was identifying markers of systolic and diastolic dysfunction in the echocardiogram, we assessed the classification performance of guideline derived cut-points for individual echocardiographic parameters in the data set.

We tested 80 patients with HFpEF all of whom were correctly identified as HF by the AI model. The clinical scores based on 2D Echocardiography and NT pro-BNP levels identified these 80 patients as HFpEF. The 50 controls were identified as normal by 2D Echocardiography and NT pro-BNP levels. Out of these 50, the AI model re-classified 24 patients as HF. Out of these, 8 controls re classified as HF by the AI model presented to the outpatient department with features of HF.

### Clinical Results

The mean age in the controls identified as normal by the AI model was 56.8±9 years and mean age in the controls identified as HF by the AI model was 52.7±12 years. There were 41 males in the control group. There were 30 (61.2%) hypertensives and 27 (55%) diabetics.

The 2 D Echocardiography showed the difference in LVEF between the controls identified as normal by the AI model (Mean=59.16, SD=0.27) was significantly greater than the LVEF of controls re classified as HF by the AI model (Mean=58.38 SD=0.30), p=0.05.

Their repeat 2D Echocardiography were not different from their baseline findings. The NT pro-BNP was significantly higher than baseline (812±12 pg/ml ; 64.2±35.7 pg/ml p<0.001).

The proportion of the AI model identifying controls as normal in control group was 0.51 (95% CI: 0.37, 0.64). The proportion of the AI model re classifying controls as HF in control group is 0.49 (95% CI: 0.36, 0.63).

The entire cohort of HFpEF patients (n=80) and controls was followed over 14 months. The overall all-cause mortality was 2 in the HFpEF group. The re-admission for heart failure occurred between 1 month to 2 years. There were 55 re-admissions for heart failure. The number and frequency of re-admissions in patients with decreased relative wall thickness (RWT) was marginally greater as compared to those HFpEF patients with preserved RWT (p=0.3) [Figure 1].

**Figure 1:**
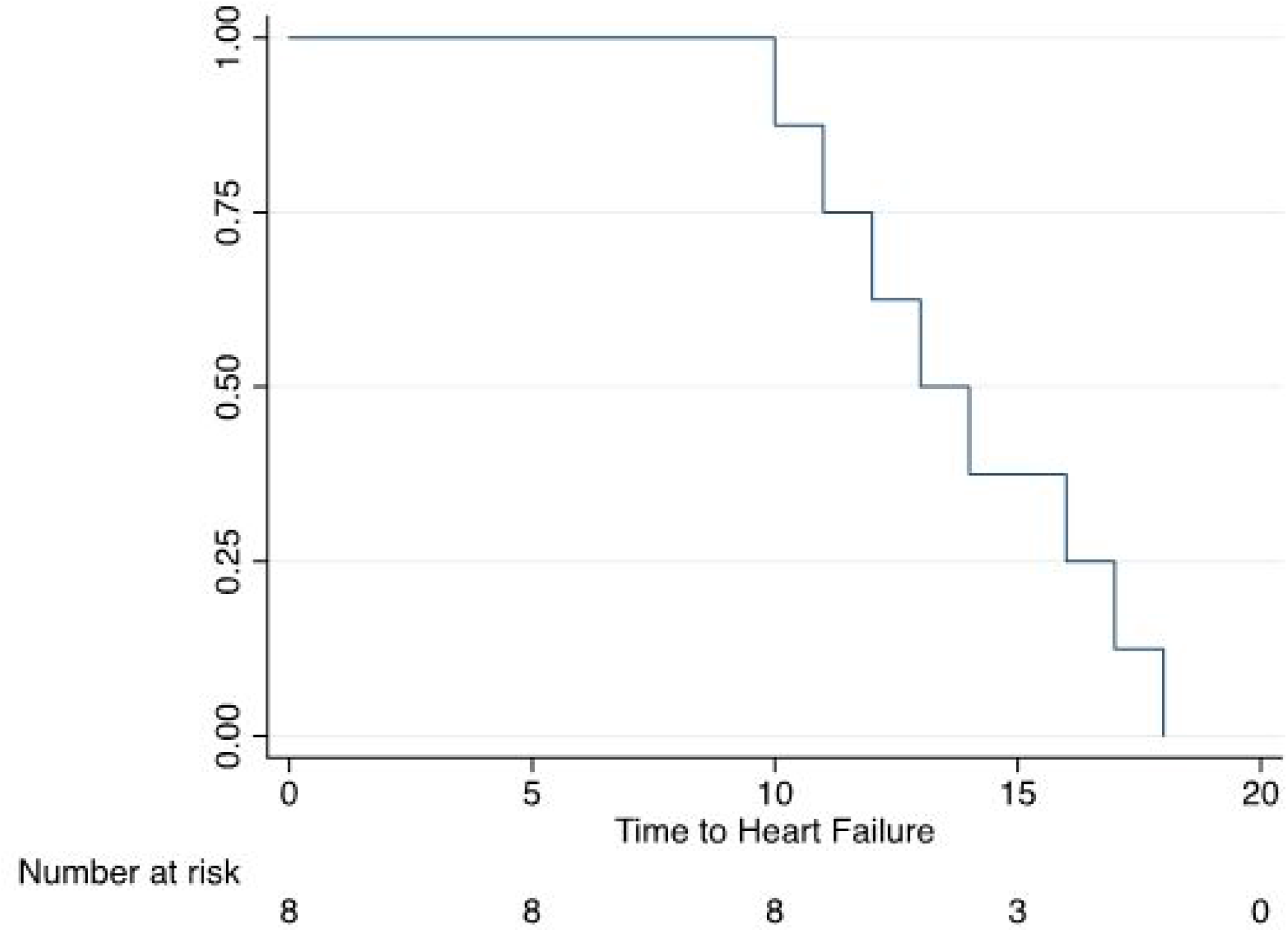
Kaplan-Meier plot of readmission during the follow-up period. Two lines represent relative wall thickness (RWT ≥ 0.34 and RWT< 0.34) groups. Shaded area represents the 95% confidence interval. The numbers at risk at the beginning of the follow-up time were n = 57 in RWT ≥ 0.34 group and n = 74 in RWT< 0.34 group. Readmissions were 23 and 32 in the 2 groups, respectively. Log-rank test P=0.3 for comparison between the 2 groups.

Out of the controls re classified by the AI model (n=24), there were 8 patients who presented with heart failure symptoms and raised NT pro-BNP levels [Figure 2]. The Kaplan-Meir plot of time to heart failure in these 8 patients is given in Figure 2.

**Figure 2:**
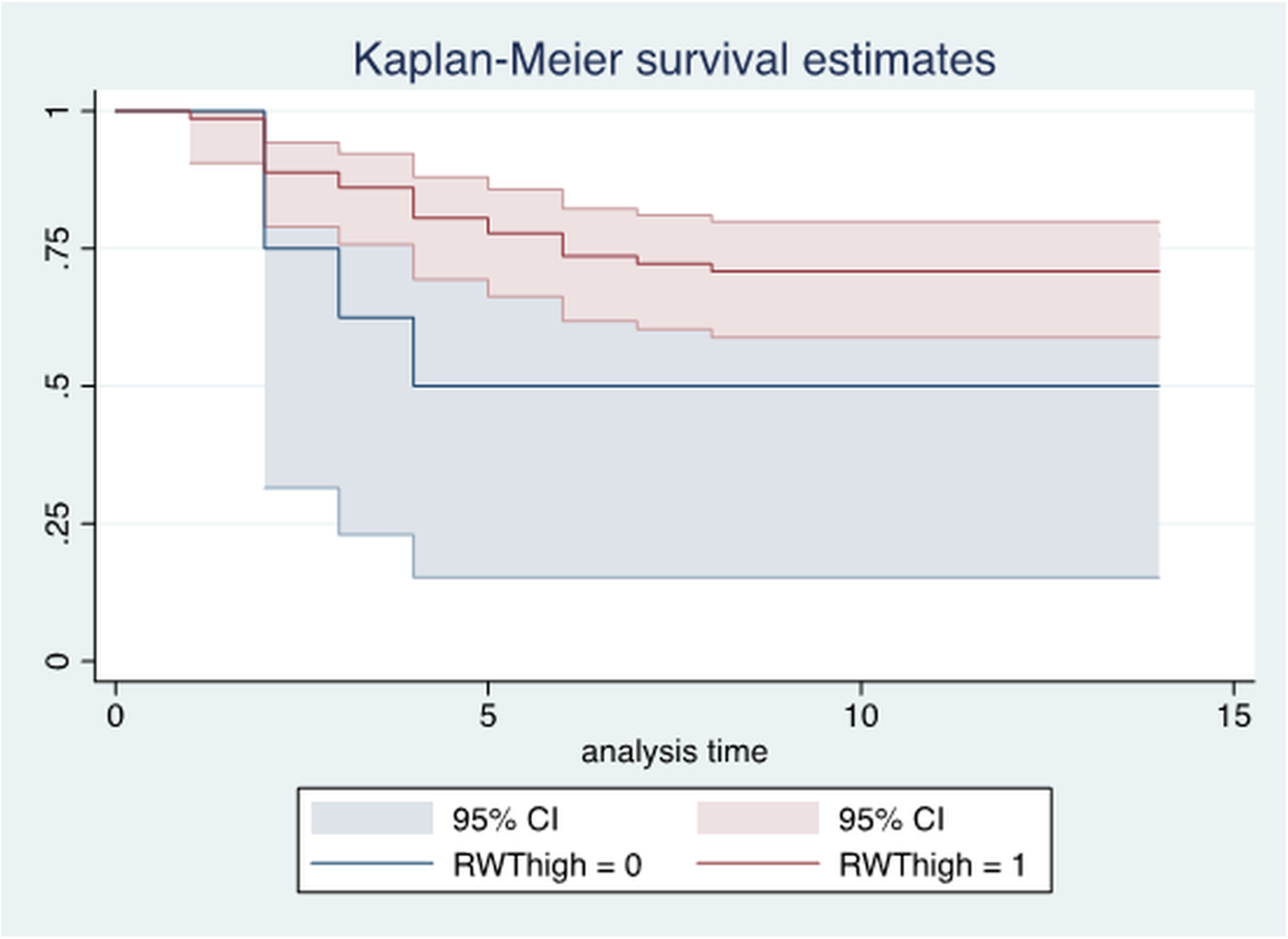
Kaplan-Meir plot of time to heart failure in 8 control patients who developed heart failure in the follow up period.

The remaining controls identified as normal (n=26) continued to be asymptomatic.

## Discussion

Ackerman et al have developed and validated a novel AI model that, used only a single A4C video clip, and demonstrated excellent ability to distinguish between patients with and without HFpEF and successfully identified patients with worse 5-year survival [6]. The ability to automatically detect HFpEF with limited clinical information has important practical use, especially in centres where a detailed echocardiographic assessment may not be feasible. The resulting misdiagnosis may prevent patient access to timely pharmacological therapy. Their model developed on a single routinely acquired video clip, demonstrated high classification accuracy consistent with comprehensive clinical and echocardiographic assessment.

The prevalence of nondiagnostic outcomes using existing clinical and Echocardiographic guidelines which include the often-tedious diastolic assessment in HFpEF are widely reported.[7,11,12-14] The performance of such methods varies considerably,[7-10] but can be excellent in some tertiary centers. In complex clinical cases, while there exists guidelines for estimating filling pressure when echocardiographic signals are difficult to interpret (atrial fibrillation [9], the assessment is often avoided entirely.

Ackerman’s AI HFpEF model retuned fewer non-diagnostic outputs, successfully reclassifying almost 75% of those who would be non-diagnostic according to the HFA-PEFF or H2FPEF scores. Further, the model identified those with increased risk of mortality, making it an invaluable addition to clinical practice; particularly in those who would otherwise be indeterminate. As it identified those with an increased risk of mortality, it may help in a higher proportion of patients being managed correctly.

Technological advances using AI provide increased capacity to identify information, which is not readily observed normally. This may occur at the expense of interpretability.

Our model helped classify HFpEF and no HFpEF with 100% sensitivity, it re classified 24 patients as HFpEF from the control group.

Comparison of cases (correctly classified) and controls (non-classified) highlights that the model has excellent sensitivity and specificity. The re classified controls might represent a cohort demonstrating provokable increases in filling pressure, or latent heart failure.

At 2-year follow-up, it was noted that 8 out of the re classified controls presented with symptoms of heart failure with elevated biomarkers (NT pro-BNP). Although long term follow-up is required to understand if the model performance in assessing HF outcomes, it helps identify those patients with subclinical disease who may require closer scrutiny and initiation of early therapy.

Further research is required to understand whether the AI model’s capability to accurately classify even subclinical heart failure will translate to clinical endpoints of reduction in hospitalizations and death.

### Study Limitations

This is a preliminary work in progress report. Therefore, a study sample bias cannot be ruled out.

## Conclusions

We have used a novel AI based model based on only a single routinely acquired TTE video clip of the A4C to accurately define HFpEF with a sensitivity of 100% and specificity of 61.9%. It also has the capability to detect subclinical heart failure in asymptomatic controls with normal NT pro-BNP levels, identifying a cohort who require more stringent observation and early treatment.

## Data Availability

DATA WILL BE AVAILABLE ON REQUEST TO EITHER AUTHOR

